# A Plantar Surface Shear Strain Methodology Utilising Digital Image Correlation

**DOI:** 10.1101/2022.06.22.22276762

**Authors:** Sarah R. Crossland, Heidi J. Siddle, Peter Culmer, Claire L. Brockett

## Abstract

The increase in the global diabetic population is leading to an increase in associated complications such as diabetic foot ulceration (DFU), associated amputations, morbidity, which substantial treatment costs. Early identification of DFU risk is therefore of great benefit. International guidelines recommend off-loading is the most important intervention for healing and prevention of DFU, with current research focused on pressure measurement techniques. The contribution of strain to DFU formation is not well understood due to challenges in measurement. The limited data available in the literature suggest that plantar strain is involved in ulcer formation. As a consequence, there is a need for plantar strain measurement systems to advance understanding and inform clinical treatment. A method was developed to determine plantar strain based on a Digital Image Correlation (DIC) approach. A speckle pattern is applied to the plantar aspect of the foot using a low ink transference method. A raised walkway with transparent panels is combined with a calibrated camera to capture images of the plantar aspect throughout a single stance phase. Plantar strain is then determined using 2D DIC and custom analysis summarises these data into clinically relevant metrics. A feasibility study involving six healthy participants was used to assess the efficacy of this new technique. The feasibility study successfully captured plantar surface strain characteristics continuously throughout the stance phase for all participants. Peak mean and averaged mean strains varied in location between participants when mapped into anatomical regions of plantar interest, ranging from the calcaneus to the metatarsal heads and hallux. This method provides the ability to measure plantar skin strain for use in both research and clinical environments. It has the potential to inform improved understanding of the role of strain in DFU formation. Further studies using this technique can support these ambitions and help differentiate between healthy and abnormal plantar strain regimes.

## Background

Approximately 7% of the UK population have diabetes [1], with this population alone predicted to increase from 4.8 million today, to to 5.5 million by 2030, and of which 1 million are currently thought to be undiagnosed [2, 3]. Associated diabetic foot ulceration (DFU) affects approximately 34% people with diabetes [4], with an expected five year mortality rate of 40% following the development of a DFU [5], which rises up towards 79% following a diabetes related amputation is required [6, 7]. Diabetic foot care accounts for £1 in every £140 spent in NHS England [8], with an upper estimate of £962 Million spent annually on DFU in 2014/15 [9]. The need to improve diabetic foot treatment pathways is clear.

Current interventions for the at-risk diabetic foot are centred on prevention and wound treatment [10], with pressure redistribution being considered the most important intervention [11, 12]. The key clinical challenge is early identification of at-risk sites prior to ulceration developing, allowing preventative action. Plantar ulceration accounts for 48% of all DFU and is the focus for most analysis techniques [13]. Both plantar pressure and shear strain are understood to play a part in the aetiology of diabetic ulcers, thus providing the opportunity to inform prophylactic or reactive treatment options [14]. Unfortunately, current technology is limited to measurement of plantar pressure and excludes shear stress and/or strain. This restricts clinical utility because pressure alone is not a good predictor of ulcer formation [15]. Figure 1 shows the potential role strain analysis of the plantar aspect could contribute to risk assessing the diabetic foot prior to ulcer formation.

**Figure 1.**
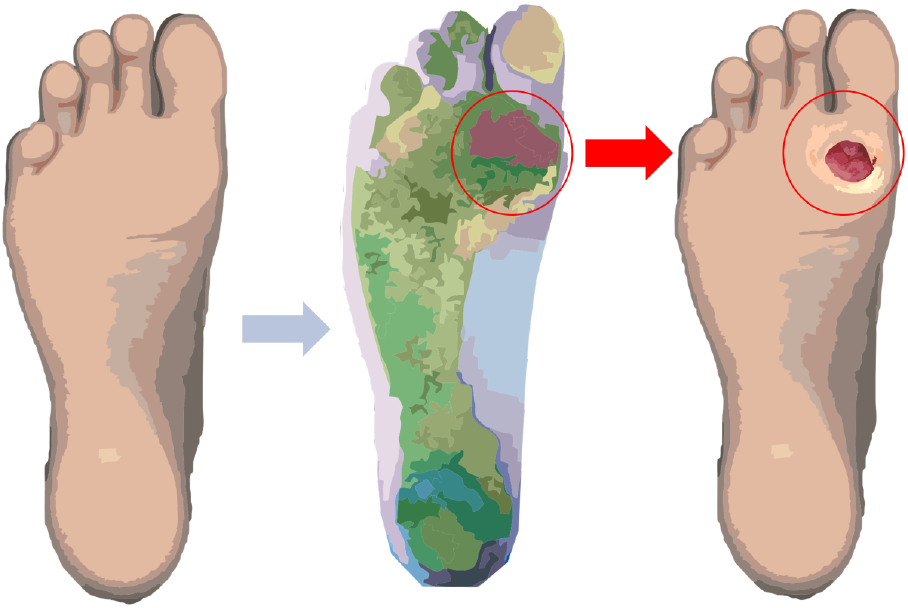
Schematic showing how plantar surface strain analysis could be used to assess diabetic ulceration risk site prior to ulcer formation.

Clinical pressure assessment can be undertaken using a wide variety of methods ranging from carbon transfer foot impression sheets for quick, low cost, pressure information, to pressure plates and insole sensors providing digitised pressure data [16]. Comparatively, plantar strain sensing technology development is in its infancy with only a range of early prototypes reported and which focus on evaluating the efficacy of different sensing approaches, including magneto-resistors, strain-gauges and capacitive sensors through to piezoelectric materials [17, 18]. Most of these methods are developed for research use and there are currently a lack of viable commercial options for clinical use [19][14]. Choosing an appropriate technology for future use in clinical settings requires consideration of factors including cost, space and time requirements, in addition to the core ability to generate clinically relevant measures. In this respect, imaging techniques have significant potential, with growing interest in analysis of static images of the foot to detect underlying abnormalities [20, 21]. Our interests are focused on measuring the dynamic aspects of strain, as it develops across the plantar aspect during gait, through the use of accessible imaging techniques.

Digital Image Correlation (DIC) is an image-based strain measurement technique. It operates by tracking positional changes of points on the patterned surface of a material as load is applied, comparing image frame(s) with a reference to determine the strain across the surface [22]. DIC emerged as a technique to track the strain of materials and started translating to biomaterials in the 1990s, with regular application after the turn of the century [23], and with increasing recent use in the field of soft materials. The use of DIC to examine strain properties within biological tissue studies is a growing field [23, 24], with in-vivo tissue studies emerging as a particular focus[25]. A key challenge with soft tissues is applying an appropriate pattern to support reliable DIC analysis. Patterns should be stochastic in nature, with good adherence to the material and clear contrast to the base material to allow for consistent pattern tracking [26, 27]. Understanding the surface strains undertaken by the skin during interactions of daily living is a pertinent topic for many applications within the field of healthcare. The foot’s plantar aspect is regularly in contact with external surfaces during gait, providing an ideal measurement opportunity to investigate its strain characteristics during loading.

Liu et al. [28] measured strain at the fingertip using DIC by imaging through a transparent glass contact surface. Similar approaches of using glass contact surfaces have been extended to observe the foot’s shape during ambulation [29]. Ito et al. [30] employed this approach with 3D DIC analysis to further investigate the feasibility of palmar and plantar surface strain tracking under laboratory conditions. These studies highlight the potential of DIC as a promising application for analysing plantar strain and informing DFU treatment. However, the methods have been developed for research purposes, requiring complex equipment and calibration which may prohibit use in a clinical environment. Typically they employ spray application of a speckle pattern onto the foot’s surface for DIC tracking. This is challenging to perform in clean clinical environments and reduces the reproducibility of coverage [27]. While laboratory studies provide a wealth of information, ensuring the technique has potential for translation to clinical utility is important.

The aim of this work is to develop a clinically suitable method to employ DIC for the measurement of plantar strain and use this to advance understanding in DFU formation. In the Methods section we first describe development of a measurement technique based on 2D DIC analysis. We then report a feasibility study used to evaluate the efficacy of the technique. Outcomes of the study are reported in the Results section employing analyses to derive a range of clinically-focused outcome measures. The paper concludes with a discussion on the advances made in this work, its relevance to research and clinical practice, and future development of the technique.

## Methods

Our approach was developed through close clinical guidance to determine a set of over-arching clinical needs. These were then used to develop and refine a concept based on technical requirements for best practice in DIC imaging and analysis.

### Clinical Needs

- Capture a single complete phase of stance during walking at a self-selected normal walking speed.
- Provide progressive tracking across the entirety of stance phase from heel strike to terminal stance.
- Preparation of the foot for imaging should be a quick procedure taking less than five minutes.
- The measurement area should be large enough to allow the entire plantar aspect to be imaged (over 95% of foot length distribution [31]) with a margin that allows for the varied placement due to stride length on to the plate.
- The method and output measures should be repeatable and linked to clinically relevant metrics.

### Concept and System Overview

A concept was developed to satisfy the clinical needs, using methods informed by the literature. Our approach, shown in Figure 2, centres on an elevated walkway within which a reinforced glass visualisation plate is embedded. The glass plate represents the target measurement area upon which the foot is placed during gait. Preparation involves patterning the plantar aspect of the foot with a speckle pattern. A high resolution camera is then used to capture a stream of images of the patterned foot throughout the stance phase. Images are processed using 2D DIC to determine strain across the plantar aspect during contact with the glass plate and subsequent analysis to determine clinically relevant descriptors related to the foot anatomy. The following sections describe the development of each of these aspects prior to evaluation in a feasibility study.

**Figure 2.**
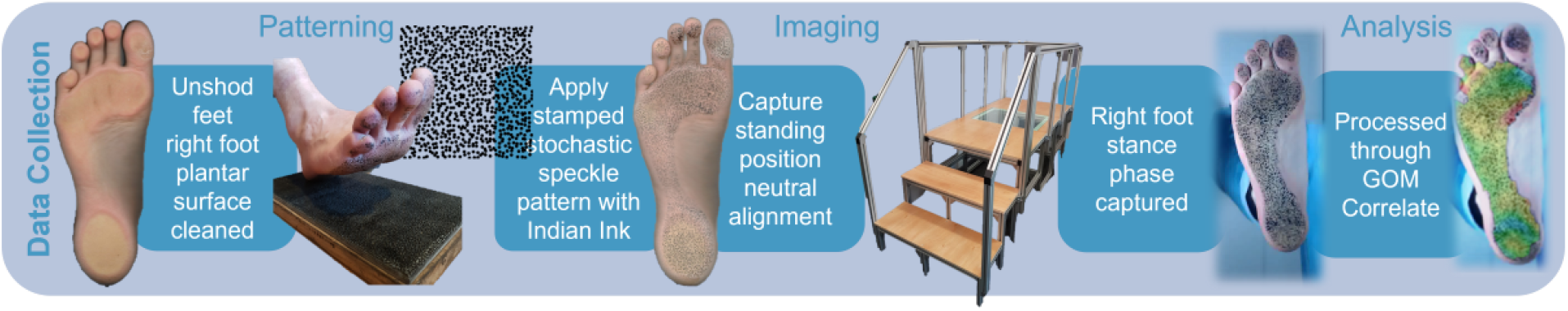
Visualisation of the data collection process for plantar surface strain assessment.

### Platform Development

The platform is designed to enable 2D DIC analysis of the plantar aspect of the foot, which enables a single camera configuration, in contrast to use of 3D DIC methods (e.g. [29, 30] which require more complex multi-camera setups). To achieve successful 2D DIC the target object should remain planar during the trial otherwise corresponding errors in measurement must be considered [26]. Our platform therefore focuses data capture on the contact phases of gait to ensure planar conditions are achieved.

Ideally the target region should almost fill the imaging field of view [26]. For a moving target, such as the plantar aspect of the foot during gait, the field of view may be slightly larger to ensure it is easy to correctly position the foot on entering stance. An appropriate depth of field to allow the target to remain in focus during the trial is also required [26]. To meet the system requirements a 2 × 1 m walkway was created with an integrated 0.8 × 0.6 m reinforced glass plate to capture plantar stance images. Half the glass plate was set as the field of view, with the plate created larger than required for single foot analysis to consider future use for imaging of either foot.

Selection of a camera and lens should allow the user to obtain the desired field of view, depth of field, frame rate and spatial resolution for clear imaging [26]. A frame rate should be determined that reduces the chance of large displacements taking place unrecorded between frames [26]. A single high-resolution webcam (1080 × 720p) operating at 30 fps was situated beneath the glass plate to meet these needs. The camera was positioned to maximise the field of view across the measurement area and provide a narrow depth of field centred on the top surface of the glass plate.

Diffuse lighting was directed at the underside of the glass plate to enhance speckle-skin contrast without introducing glare to ensure consistent pattern tracking [26]. During testing, room lighting was dimmed to minimise specular reflection.

The imaging system was calibrated using a multi-image chequerboard process to determine the real world image size for subsequent analysis (MATLAB [R2021a]). The calibration process (termed intrinsic calibration) determines and corrects for lens distortion for a single camera with a standard lens by transforming the real world points in x,y,z using a matrix of the determined camera parameters [32–34].

### Pattern Application

The quality of DIC measurement relies upon application of a repeatable stochastic speckle pattern, for which speckle movement can be tracked and post analysis performed [26]. The selected pattern must consider speckle-pixel ratio, variance in pattern and pattern density to ensure it can be appropriately tracked [26].

To pattern the plantar aspect of the foot, a range of application methods were trialled, with selection criteria being 1) ease and speed of application to different foot shapes, 2) pattern quality after application 3) repeatability of the method. Methods included hand patterning, spray patterning (commonly optimised for use in DIC [23, 24]) and temporary tattoo transfers (as employed in wearable electronic sensors [35]). A low ink transference rubber stamping method was selected because it offers quick, consistent and high quality patterning whilst being a relatively clean process compared to spraying techniques. This comprises of a 300 × 150 mm rubber stamp with a wooden base to prevent bowing during full weight transference onto the stamp. Indian ink was applied, as it is skin safe and has a low allergenic response, with a rubber roller to reduce the quantity of ink transferred to prevent pooling on the stamp. The stochastic speckle pattern used was computer generated using Correlated Solutions Speckle Generator [v1.0.5].

Patterns of varying speckle diameter and pattern density were trialled from 0.75 - 2 mm based on recommendations from best practice [26, 27], with a standardised pattern variation set at 75% for a stochastic distribution. Trialled speckle diameters were selected to meet machining requirements for stamp creation. To optimise the pattern for the output, 55 × 55 mm skin representative samples Smooth-On Ecoflex™ 00-30 with an embedded spandex layer were patterned to undergo strain testing. Tensile testing studies were conducted to expected peak mechanical plantar skin strains of 0.55, using an Instron^®^ 5943. This strain was chosen in line with the mechanical properties of excised healthy and diabetic cadaveric plantar skin samples [36, 37]. This given strain should exceed peaks seen for surface strains of in-vivo tissue sufficiently to assess pattern tracking consistency. GOM Correlate 2020 [v2.0.1] was used to track pattern quality throughout the extension to peak strain. We considered the optimal pattern would be able to track to the peak strain implemented with a loss of 5% or fewer tracked points, as pattern degradation should be minimised to reduce decorrelation during analysis [26]. A 1.25 mm speckle with 75% density and variation of pattern, see Figure 2, was found to be optimal for the output and used to form the rubber stamp. Simplification of the speckle pattern application method was required to ensure ease of translation to potential clinical environments for DIC.

### DIC Plantar Strain Analysis

A commericial DIC software package was used to perform the DIC post analysis of images and thus form a time series of strain maps for the plantar aspect of the foot (GOM Correlate). First, high resolution surface strain maps were generated for the stance phase. Strain maps were then generated for each image frame captured, to provide a detailed picture of the variance of surface strain throughout individual phases of stance with respect to a neutral standing position. An equidistant point spread was applied across the surface of the strain maps to derive strain and positioning data sets for post-processing. Strain data within a 3 *σ* distribution from the mean were selected for export, to minimise outliers and artefacts caused by out of plane motion from skewing the data distribution and display in the generated quiver and heatmap plots, as shown in Figure 3.

**Figure 3.**
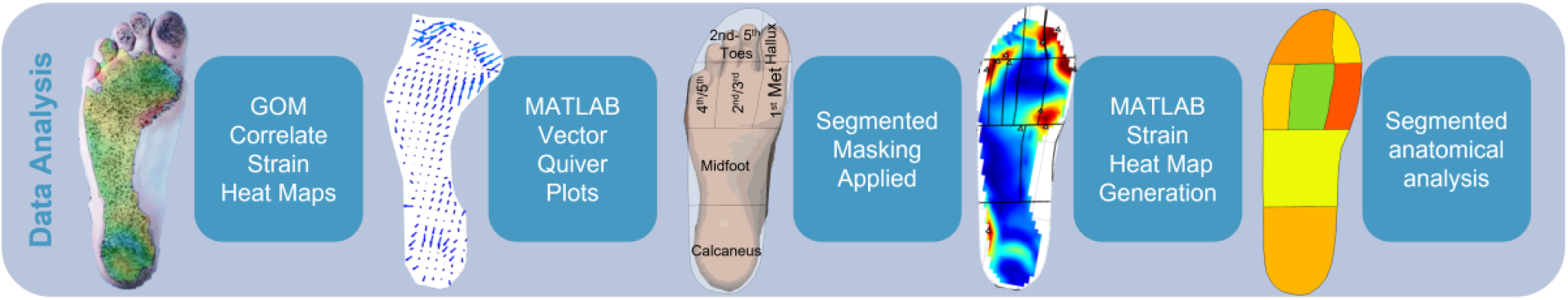
Visualisation of the data analysis process for plantar surface strain assessment.

### Regional Summary Analysis

The exported strain map data determined through DIC were then post-processed for visualisation and to derive summary metrics using custom scripts in mathematical analysis software (MATLAB). A seven region mask segmented with continual borders was linearly transformed and overlaid onto the reference images by a qualified orthotist using anatomical landmarks, as shown in Figure 3. Segmented masking of the plantar aspect enabled comparative analysis of strain across the foot and particularly in areas at high-risk of ulceration. 75% of plantar DFU occurs at the forefoot regions [38], with a near even concentration between the toes and metatarsal regions [13]. The defined regions were determined based on these known ulceration prevalent anatomical regions, in known regions of interest for DFU [39], to define regionalised surface strain data. This is consistent with similar plantar masks used to report pressure in other systems such as Novel’s ^®^ Automask which divides the foot into anatomically significant segments for zonal analysis [40]. The anatomical regions are segmented into the hindfoot, midfoot, individual metatarsal heads, hallux, second toe and combined third to fifth toes [40]. Initial masking was defined to cover these regions and divide the midfoot, which was then simplified to the chosen seven region mask when analysing data output to consider difficulty in mask tracking during gait. Through this segmented approach the highest recorded (peak) mean strain seen in one frame within stance phase and the averaged mean strain across all recorded frames in each region were determined.

### Participant Study

A feasibility study was conducted to evaluate the efficacy of the proposed methodology. In particular it aimed to assess the ability of the experimental set-up to meet the requirements for effective data collection, analysis and the clinical needs outlined. The study was designed to use the DIC methodology to collect right foot single stance phase surface strain analysis of the plantar aspect of a subset of six non-diabetic participants, see Table. 1. Ethics were obtained through the University of Leeds Engineering and Physical Sciences joint Faculty Research Ethics Committee (LTMECH-001) to meet the requirements of testing. The experimental protocol for data collection is shown in Figure 2.

**Table 1.** Six participant profile data.

| Gender | Weight | Shoe Size |
| --- | --- | --- |
| Male | 61.6 - 96.9 kg | 8-12 UK Male |
| Female |  | 5.5-7 UK Male |

Each participant underwent the patterning process, which required the right foot to be wiped clean and then stamped using the low ink transference stamping method. Camera calibration. was performed prior to each participant using the system. A reference image was taken to provide a baseline for the surface strain of the skin to be compared to following standing and walking. This image was captured when a full weighted neutral stationary standing position was adopted on the glass plate of the walkway. Following this a single step was taken from a standing start on to the glass at a self-selected normal walking speed and images were captured from the camera at 30 Hz.

## Results

All six participants completed the data collection process successfully. Figure 4 shows an example of a multiframe heat map output generated by the DIC analysis software (GOM Correlate) throughout stance comparative to a neutral standing position. Figure 5 shows a single frame of data after post-processing to produce a strain quiver plot and segmented heatmap for region specific analysis. A variation in the number of frames to complete stance phase was seen across the participants due to variations in the biomechanics and speed of gait. In general, the magnitudes and direction of strain were observed to vary greatly across the aspect of the foot and throughout gait for each participant.

**Figure 4.**
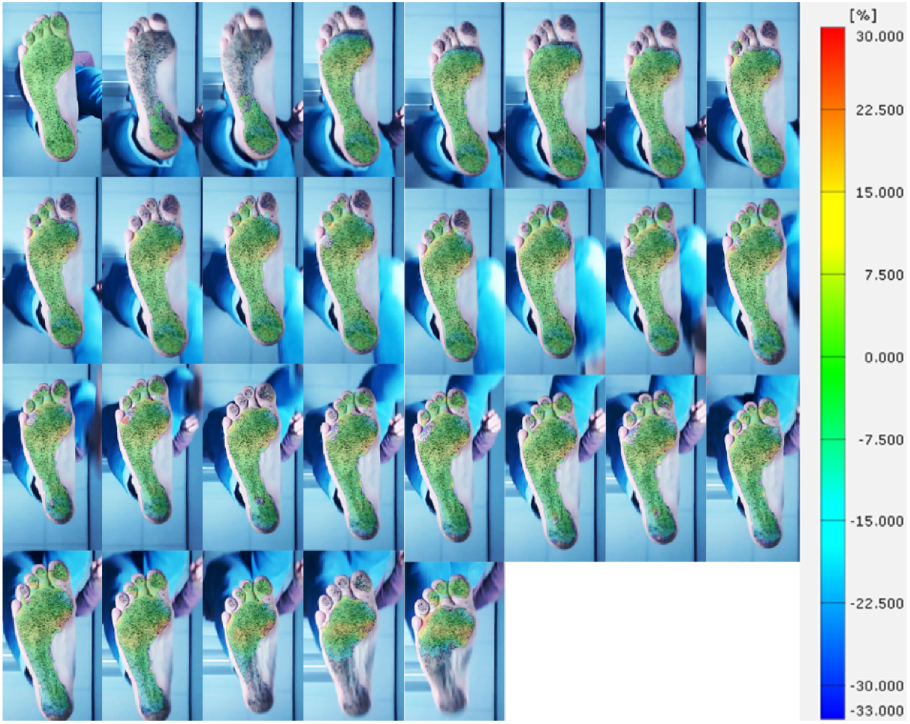
Example surface strain GOM Correlate heat map comparative to standing throughout stance.

**Figure 5.**
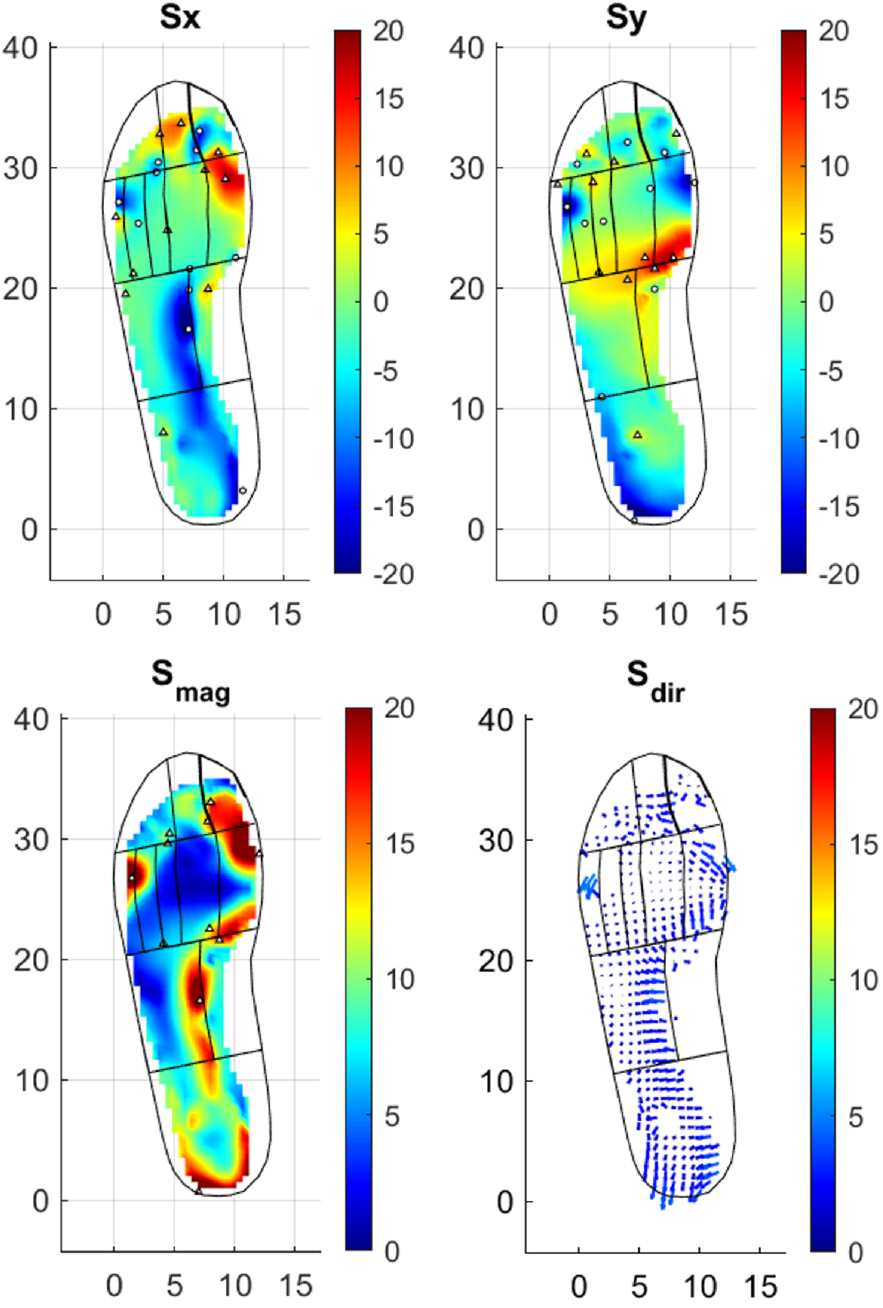
Example surface strain of a MATLAB generated heat map from one frame of stance. Showing strain independently in x and y, the combined magnitude and comparative quiver plot.

The regional analysis of mean strain and standard deviation for each frame of stance phase per participant was conducted. From this, the peak mean value that is seen in each anatomical region during stance was determined. Figure. 6 shows a graphical representation of the distribution of peak mean strain and standard deviation per region for each participant. Across the participants a relatively large standard deviation can be seen around the mean, showing the substantial spread of strains seen within localised regions during stance. Comparatively the range of peak mean strains is relatively consistent between the group. Although the distribution of strain is varied per participant a pattern can be seen in the colour spread with higher mean values often focused around the calcaneus and forefoot with lower means seen in the midfoot.

**Figure 6.**
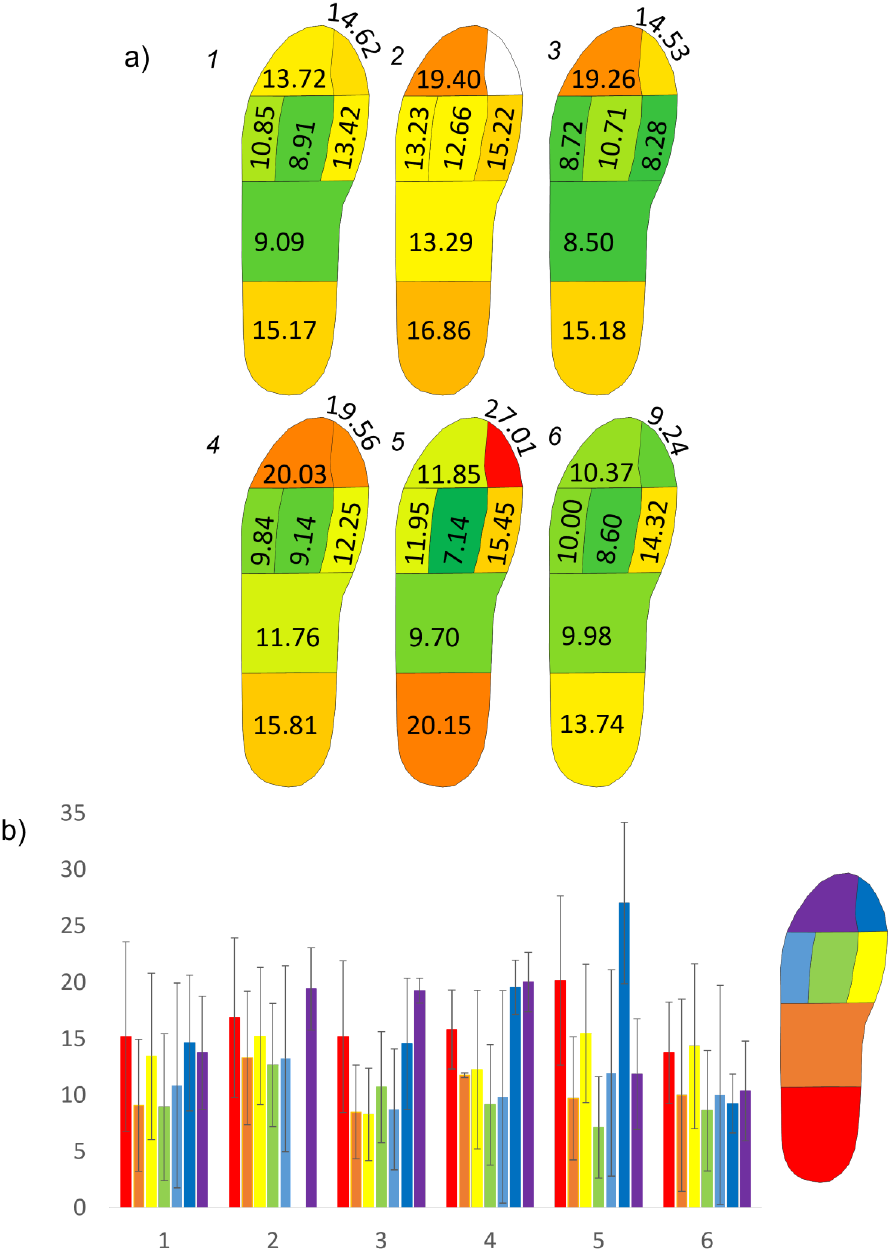
a) Peak mean values foot maps with a colour distribution overlay showing regions of highest (red) and lowest (green) strains. b) Peak mean strain per region and standard deviation throughout all frames of stance per participant.

Figure. 7 shows the averaged mean strains occurring in each region throughout the entirety of stance. The segmented foot maps colour distribution shows in which regions the highest and lowest strain occurs respectively across all participants. The range of the averaged means is relatively low, but the distribution of strain across the plantar aspect between each participants is again pronounced. Similar participant strain distributions at the forefoot and calcaneus are seen between the peak mean and averaged means. It can be noted a few participants displayed moderate to high midfoot means comparative to the entire plantar aspect.

**Figure 7.**
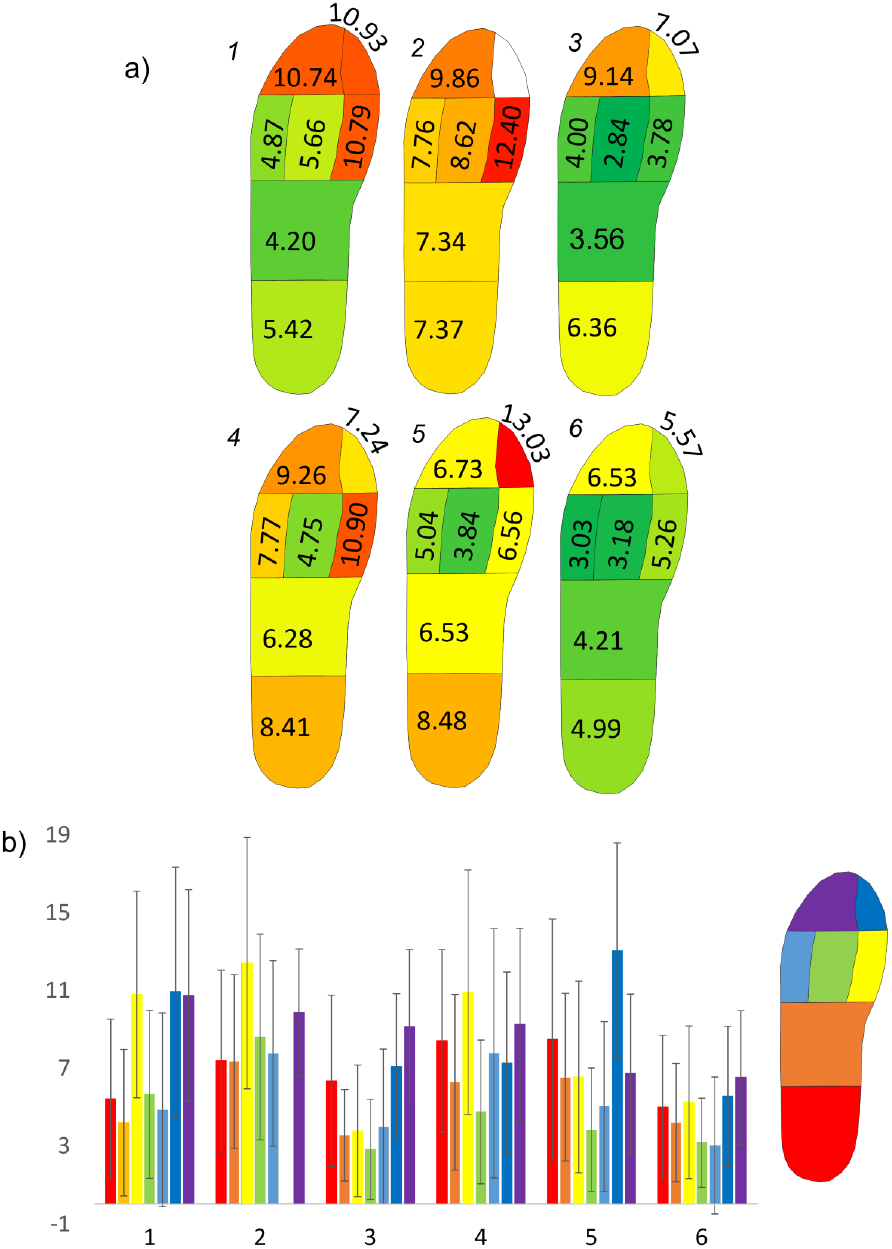
a) Averaged mean values foot maps with a colour distribution overlay showing regions of highest (red) and lowest (green) strains. b) Averaged mean strain per region and standard deviation throughout all frames of stance per participant.

Variation in peak mean and averaged mean patterns for each participant reflects the variation in foot contact and loading during stance seen in the participant trials. Typical biomechanical positioning of the foot throughout stance is considered as progressing from the lateral hindfoot through to the medial forefoot [41], this is reflected in the higher strain regions for both peak and mean strain values generally across the participant group. Participant 2 did not register any DIC tracking in the hallux segment, as seen in Figures. 6,7. This indicates a potential lack of contact during stance phase within the plane of tracking.

## Discussion

Our principle aim in this work was to develop a robust methodology to assess plantar surface skin strain in gait using DIC. This was developed and successfully trialled through a six-participant study of the stance phase in gait. Our emphasis was on developing methods appropriate for use in clinical practice. Using an ink stamp for the speckle pattern application allows for a quick and uniform patterning of the entire plantar aspect. The pattern is pre-determined and consistent which increases the reproducible nature of the method comparative to traditional pattern spraying techniques. It also affords comparatively clean application, which would be beneficial within both clinical and research environments. The walkway, camera and lighting set-up offered a regulated environment in which to conduct repeated studies under the same environmental conditions to meet the system requirements for successful DIC. By operating a 2D data capture method, there is a reduction in equipment required, which costs less and removes the need to run cameras in stereo. Removing the complex set-up 3D DIC requires for the stereo process, saves time and associated errors, making it much more accessible in the clinical environment.

Alongside the measurement methodology, it is equally important to consider the development of clinically relevant output measures. Whilst commercial DIC packages (e.g. GOM Correlate) provide a generalised overview of the strain, exporting the data for custom post-processing and visualisation extends the options for foot-specific clinically-focused analysis. This can help support further understanding of the properties of surface skin tribology to understand the contribution of strain to DFU formation. Applying the 3*σ* exclusion criteria to the data to exclude outliers brought about via out of plane motion being tracked, reduces the error inherent in 2D DIC processes in the data presented. Using segmented masking across the plantar aspect of the foot provides the ability to consolidate the wealth of temporal strain information obtained from this measurement process into specific regional plantar analyses, to work towards informing treatment approaches. This can inform both targeted analysis of anatomical locations during select phases of gait, such as during propulsion, and peak strains across the foot as a whole. Evaluating the peak mean allows us to identify the highest average strain applied to a specific region during stance phase. This will enable the researcher or clinician to assess if a participant is consistently experiencing a high strain over a particular region, indicating potential sites at increased risk of DFU. Frame by frame temporal analysis provide the opportunity to assess if these peak means align with particular phases of stance, such as at terminal stance. Comparing this to the averaged mean can provide a baseline compared to standing of the strain experienced by the same region throughout stance. Comparatively, analysis of singular strain peaks, whilst offering indication of key areas of concern, may not see sustained high strains throughout the duration of gait or beyond a singular point on the plantar aspect. With the pressure-time integral has been highlighted as important when measuring direct pressures [42]. Both mean and peak data sets should be considered together to provide a rounded picture of the strain changes during stance. These outputs offer the potential for further tribological investigation into DFU formation in specified regions using strains derived from this DIC methodology.

The feasibility study provided valuable insight into the potential clinical relevance of this method. The segmented analysis showed moderate to high values of strain within the participant group at the midfoot. The midfoot experiences a lower DFU incidence than regions such as the 1st metatarsal head [13], and has reduced ground contact especially medially due to the longitudinal arch of the foot and likely to experience lower strains. Comparing the segmented values and the generated strain heat maps, it does not appear that high strains develop in the midfoot region. This implies that the mask, defined on the static reference image does not track as accurately on the corresponding frames during the stance phase of gait. It is likely that tissue strain and deformation of the foot can result in discrepancies with respect to the mask, for example the calcaneus may be represented in the midfoot region. This highlights a potential limitation of using a static mask during this part of the analysis process. Further development to ensure an adequate tracking mask is applied to each frame is necessary; for example adapting the masking for each frame is possible with the commercial Novel ^®^Automask system and may provide a potential solution.

The principal limitation of this method is the inherent unshod nature of the foot meaning it cannot accurately recreate the shod environment. This aligns the method with pressure plate studies which are also conducted unshod and regularly used within clinical environments. However walking in footwear accounts for most steps taken daily and should be considered when determining how plantar tribology contributes to DFU formation. By undertaking 2D plantar DIC we can gain an understanding of the surface skin interaction with a set opposing surface, which can be used to inform further tribological study of the plantar foot during gait. However, the errors which arise from out of plane motion vary per participant and must be adjusted by blanket data exclusion criteria, as it is impossible to account for the in errors arising from differing gait. 3D DIC is advantageous in this respect and offers the potential for dorsal strain mapping but requires assessment within footwear.

Within the six participant study a small number of frames were unable to be tracked during the midportion of stance phase. It was potentially due to the poor image quality of certain frames as a result of the camera lagging during operation. It was considered that sufficient information is provided on the remaining frames for this not to hinder the surface strain outputs seen in the regions analysis. An improved imaging system with a higher frame rate can readily improve this process without significant change to the technique. Considering the potential transference to clinical utility, patterning using low ink transfer stamping is suited to a time pressured and clean clinical environment. The current walkway may be unsuitable in limited space settings but could be reduced in size once clinical efficacy is determined.

Following this developmental work, an extended participant study is warranted to further understand the data analyses required to effectively characterise the plantar skin strains and define normative ranges. Further research is required to understand the contribution of strain in the mechanical formation of DFU. These data will be used to inform tribological studies which replicate plantar interactions to assess skin degradation and deep tissue strain responses. Supporting this method with shod analysis techniques would also be beneficial and should be investigated further.

## Conclusions

Using DIC to measure plantar surface shear strain by applying a computer-generated pattern using low ink transference stamping offers a process that is quick, relatively low mess, reliable and provides repeatable patterning, through the set nature of the stamp profile. This technique, based on a 2D DIC method, allows for coherent tracking of strain on the plantar aspect of the foot during the frames of stance phase captured comparative to the reference image, as demonstrated in a six participants feasibility study. Temporal aspects of the stance phase were tracked from heel contact through to propulsive terminal stance. Compared to current in shoe shear strain sensors, this technique offers high resolution data of the plantar aspect of the foot throughout stance phase. This technique offers potential to become a clinically viable tool but further investigations are needed to support this method with more work required in understanding the role that strain plays in DFU formation.

## Data Availability

The datasets generated and/or analysed during the current study are
available on request.

## Abbreviations

DFU: Diabetic Foot Ulceration
DIC: Digital Image Correlation

## Declarations

### Ethics approval and consent to participate

Ethics were obtained via the University of Leeds Engineering and Physical Sciences joint Faculty Research Ethics Committee. Ethics Reference: LTMECH-001. Consent was obtained from participants of the feasibility study.

### Consent for Publication

Consent to use data for publication was obtained from participants in the feasibility study.

### Availability of data and materials

The datasets generated and/or analysed during the current study are available in the University of Leeds Data Repository, [doi to be provided before publication]. [43]

### Authors’ Contributions

All authors contributed to the concept and research ideas. SRC developed the methodology and collected the data. SRC and PC were involved in analysis. All authors read and approved the final manuscript.

## Acknowledgements

Andrew Stockdale, University of Leeds, must be thanked for his technical support on the fabrication of the walkway for this study. I would like to extend my thanks to Leeds Teaching Hospitals NHS Trust for allowing me to work in their Diabetic Limb Salvage Service on an honorary basis which has influenced my drivers and motivation for this study. Thanks should also be extended to my supervision team for their continual support and guidance and to EPSRC for funding this research.

